# Mucosal immunity and antibody anergy in COVID-exposed Covishield vaccinees

**DOI:** 10.1101/2022.09.06.22279625

**Authors:** Priya Kannian, Pasuvaraj Mahanathi, Arul Gracemary, Nagalingeswaran Kumarasamy, Stephen J. Challacombe

## Abstract

Knowledge is limited on mucosal immunity induction and longitudinal responses to vaccination against SARS-CoV2. Here, we determined serum/salivary antibodies and cytokines after three Covishield vaccine doses. Sera from 205 healthcare workers (HCWs) one-month after first-dose; one-, three- and six-months after second-dose; paired sera and stimulated whole mouth fluid (SWMF) from 10 HCWs one-, three- and six-months after third-dose were tested for anti-spike SARS-CoV2 antibodies by ECLIA and for cytokines by ELISA/cytokine bead arrays. One-month after second-dose, antibodies had increased significantly (6-fold) in COVID-naïve group, but declined (1.5-fold) in those previously exposed to COVID. At one-month after first-dose, IL-10 levels were statistically higher in the previously COVID-exposed group compared to COVID-naïve group (p<0.02). Breakthrough infections were 44% in COVID-naïve group, while re-infections were 27% in COVID-exposed group (p<0.02). Proinflammatory cytokines–IL- 17/IL-21 at one-month after first- and second-doses, and memory cytokines–IL-7/IL-15 at three- and six-months after second-dose were minimal. Antibodies spiked at one-month after third-dose and declined by three- and six-months after third-dose similar to post-second-dose. Paired sera and SWMF at one- and six-months after third-dose lacked adaptive immunity cytokine expression. Innate immunity cytokines (MIG, MCP-1, IL-8, TNF-α, IL-6, IL-1β) showed a declining trend in serum, but were sustained in SWMF. Thus, our findings suggest that first-dose acts as an antibody boost, while second-dose induces antibody anergy in the previously COVID-exposed group. Rapidly declining antibodies and minimal T cell cytokines raises concerns over their durability in subsequent virus exposures. Sustained innate cytokines emanating from the oral mucosa warrant further in-depth explorations.

## Introduction

Vaccines confer protection by inducing certain effector mechanisms including antibodies produced by B cells, CD8^+^ T cells and CD4^+^ T cells. The CD4^+^ helper T cells (Th cells) mediate protection through cytokines and contribute to the generation and maintenance of B cells and CD8^+^ T cells. CD4^+^ effector T cells that play a major role in vaccine induced protection include the follicular helper T cells (Tfh cells), Th17 cells and regulatory T cells (Tregs). Tfh cells are primarily positioned in the local lymph nodes where they mediate B cell activation, differentiation, somatic hypermutation and class switching. They directly control the antibody responses and mediate adjuvanticity. Th17 cells promote a local pro-inflammatory response and contribute largely to mucosal immunity. These effector T cells are controlled by the Tregs. The cytokines produced by these effector and regulatory T cells that contribute to the effectiveness of a vaccine include IL-17, IL-21, IL-10, IL-7 and IL-15 (1).

The predominantly administered COVID-19 vaccine in India is the Covishield vaccine, which is identical to Astra Zeneca’s ChAdOx1, and manufactured under licence. It is a chimpanzee adenoviral vector vaccine carrying the genetic material coding for the spike protein. Studies from India have reported that the Covishield vaccine reduces the incidence rate of infection, transmission rate, mortality rate and rate of hospitalization compared with unvaccinated controls (2-5). Although these large cohort studies have shown the effectiveness of this vaccine, there is little knowledge about the T cell response elicited. Robust T effector and memory cell responses are important for the activation of B cells and maintenance of memory B cells. Studies have demonstrated differential humoral and *in vitro* cell-mediated immune responses in COVID-naïve and COVID-exposed individuals post-vaccination with two doses of BNT162b2 vaccine and after one dose of Ad25.CoV2.S vaccine (6,7). Since these studies were performed *in vitro* upon stimulation of lymphocytes with specific peptides, the data should be interpreted cautiously, as these findings do not necessarily correlate with *in vivo* reduction of antigen burden by the innate immune response and antigen presentation to the T cells. In addition, there is very little data on the induction of mucosal immunity by Covishield vaccination.

Therefore, in this study we proposed to determine the antibody responses, and the pro-inflammatory and memory T cell responses in Covishield vaccinees in an *ex vivo* manner up to six months after the third vaccine dose, and to examine mucosal immunity in a sub-group over six months. Our findings help to elucidate the B cell and T cell responses to the Covishield vaccine up to six months after the second dose in people exposed and not exposed to COVID-19 prior to vaccination. We compared these responses in people who developed COVID-19 after vaccination with those who did not develop COVID-19. The innate and adaptive immunity cytokines have also been elucidated at one and six months after the third booster dose in paired serum and stimulated whole mouth fluid (SWMF) samples.

## Methods

### Materials and Methods

#### Patients and samples

The study was approved by the VHS-Institutional Ethics Committee (Proposal# VHS-IEC/72-2020). Healthcare workers (HCW; n=220) were recruited one month after first vaccine dose of Covishield (same as ChAdOx1 manufactured in India) after written informed consent. Blood samples were collected at the time of recruitment. Demographic details, past COVID-19 exposure and vaccination dates were collated. Subsequent longitudinal blood samples were collected on 1^st^ month, 3^rd^ month and 6^th^ month after the second dose. Longitudinal blood and SWMF samples were collected at 1^st^ month, 3^rd^ month and 6^th^ month after the third booster dose. Sera and SWMF supernatant samples were separated immediately by centrifugation (serum: 3500rpm for 25 minutes; SWMF: 1500rpm for 10 minutes) and stored at -80ºC until further use. HCWs lost to follow up after the first dose-first month samples were excluded from the study.

After exclusion, 205 HCWs were included for the testing and analyses. Additionally, SWMF samples were collected from 11 unvaccinated COVID-naïve healthy controls.

#### Anti-SARS-CoV2 spike and nucleoprotein antibodies assays

Anti-SARS-CoV2 spike and/or nucleoprotein antibodies (total Ig including IgG) were detected by electrochemiluminescence assay (ECLIA) using Cobas e411 automated analyser (Roche, Germany). Values <1 cut-off index (COI) were considered negative for the anti-nucleoprotein antibody kit. Values <1 U/ml were considered negative for the anti-spike antibody assay. Samples with values >250 U/ml were serially diluted to 1:400 with 1x Dulbecco’s phosphate buffered saline (PBS; HiMedia, India) and the final antibody concentration was determined. For SWMF samples, the total protein content (mg/dl) was determined by the pyrogallol red molybdate method (Diemnsion Xpand, Siemens Healthcare, USA) to normalize the cytokines and chemokines in the SWMF samples.

#### Cytokine assays

IL-17, IL-21 and IL-10 levels were tested at one month after vaccination, while the memory cytokines – IL-7 and IL-15 levels in the stored serum samples were tested at three and six months after the second vaccine dose by enzyme linked immunosorbent assay (ELISA), according to the manufacturer’s instructions (R&D systems, USA). Cytokine bead arrays (CBA) were performed on the serum and SWMF samples using the BD CBA flex beads as per the manufacturer’s instructions by flow cytometry (BD FACS Lyric, BD Biosciences, USA).

#### Statistical analysis

The mean, median and quartile values were calculated using Microsoft Excel. The statistical significance between the groups was calculated using the free online t-test, Anova test or Mann-Whitney U test calculators from Social Science Statistics or Vassar Stats.

## Results & Discussion

Of the 205 cases analysed in this study, 111 (54%) were COVID-naïve and 94 (46%) were COVID-exposed prior to vaccination. Among the COVID-naïve group, 78/111 (70%) developed detectable anti-SARS-CoV2 spike antibodies at one month after the first dose and 104/111 (94%) at one month after the second dose. The immunogenicity of the Covishield vaccine in our study was similar to other studies with the Astra Zeneca vaccine as well as other mRNA vaccines (8,9). The median antibody response at one month after the first dose in the COVID-exposed group (median: 10152 U/ml) was 121-fold higher than the COVID-naïve group (median: 84 U/ml; p<0.0001; Figure 1A). The antibody response in the COVID-naïve group at one month after the second dose (median: 856 U/ml) was still significantly lower than the antibody response in the COVID-exposed group at one month after the first dose (median: 10152 U/ml; p<0.0001).

**Figure 1:**
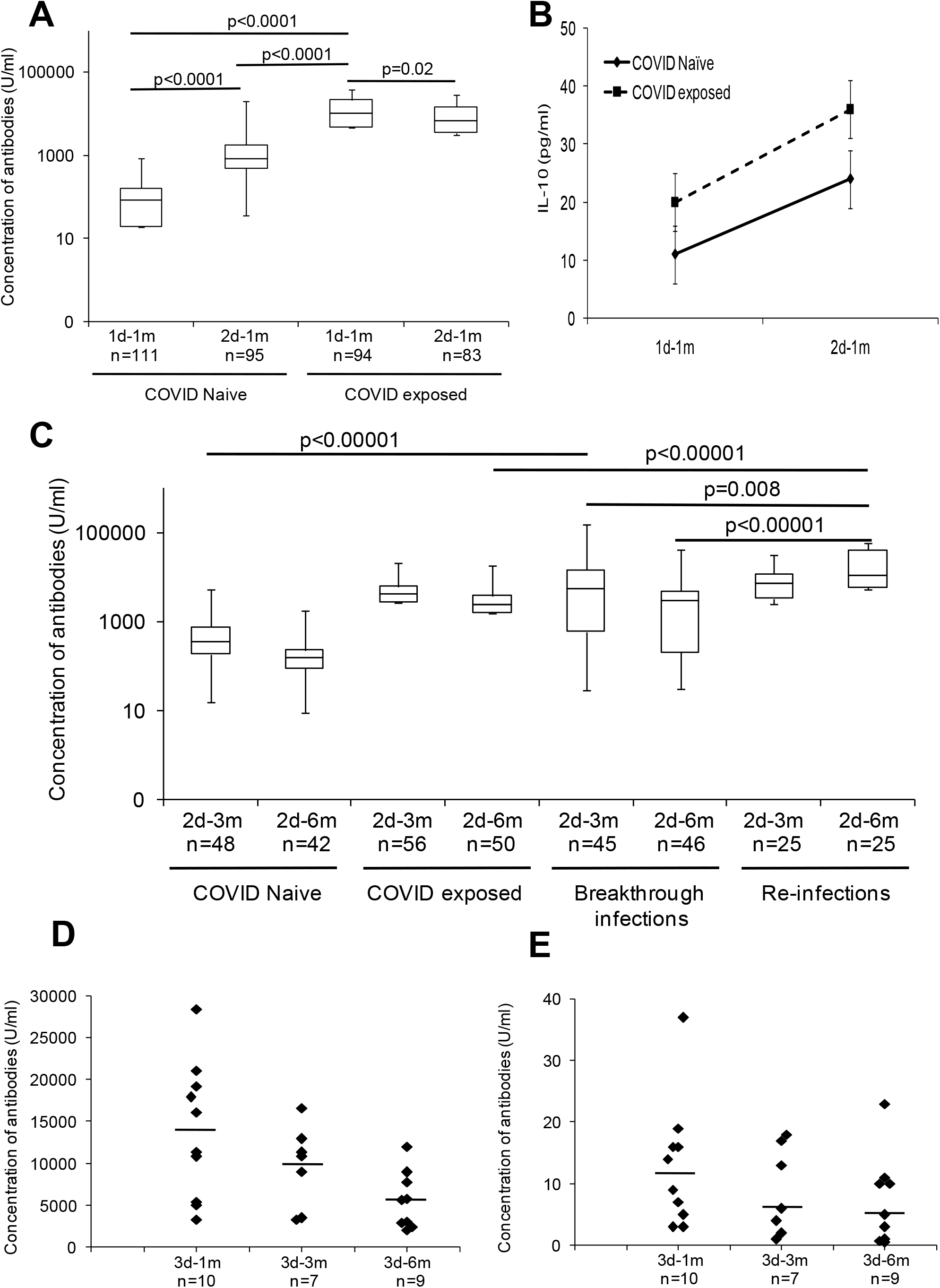
Serum anti-spike SARS-CoV2 antibodies and IL-10 levels in the COVID-naïve and COVID-exposed groups after the first (one month), second (1, 3, and 6 months) and third (1, 3, and 6 months) doses of the vaccine. n= the number of samples in each group. Y-axis denotes logarithmic concentration of the antibodies (U/ml). The box denotes median and interquartile ranges. The bottom and top whiskers denote the minimum and maximum values, respectively. The p values indicated were calculated using Mann-Whitney U test. **A: COVID-naïve and COVID-exposed groups at one month after the first and second doses of the vaccine. B: Levels of IL-10 in COVID naïve and exposed groups**. Black diamonds - COVID-naïve group (n=24); black squares - COVID-exposed group (n=36). The error bars indicate the standard deviation. 1d-1m: p=0.01, 2d-1m: p=0.02 (t-test). **C: Serum** a**nti-SARS-CoV2 spike antibody responses at three months and six months after the second dose in COVID-naïve, COVID-exposed, COVID-naïve group with breakthrough infections and COVID-exposed group with re-infections. D and E: Anti-SARS-CoV2 spike antibody responses at one, three and six months after the third booster dose in serum (D) and SWMF (E) samples**. Bars represent the median.

Thus a natural infection primes the immune response much more effectively than Covishield vaccination. However, the antibody responses elicited after two doses of BNT162b2 vaccine is proportional in both the COVID-naïve and COVID-exposed groups (6). The antibody response in the COVID-naïve group showed a statistically significant 6-fold increase at one month after the second dose (Figure 1A) compared with the first dose (p<0.0001). On the other hand, the median antibody level in the COVID-exposed group unexpectedly showed a statistically significant 1.5-fold decline at one month after the second dose (median: 6986 U/ml; Figure 1A) compared to the first dose (p=0.02).

The mean IL-10 level (regulatory cytokine) was statistically higher in the COVID-exposed group (mean: 20pg/ml) at one month after the first dose (Figure 1B) compared with the COVID-naïve group (mean: 11pg/ml; p<0.01). At one month after the second dose the IL-10 levels in the COVID-naïve group (mean: 24pg/ml) increased significantly from one month after the first dose (p<0.02); and the IL-10 levels were similar to that of the COVID-exposed group, at one month after the first dose time point, suggesting a regulatory response at the time of a second antigen challenge. Thus the antibody response elicited in the COVID-naïve group after two doses indicates a prime-boost response, while in the COVID-exposed group prior to vaccination the first vaccine dose acts as a booster and the second vaccine dose appears to induce anergy with regard to antibodies. A similar anergic effect was shown by Lozano-Ojalvo *et al* where the antibody responses in the previously infected vaccinees plateau after the first dose (6).

With regard to the question of whether the immune responses were protective against further SARS-CoV2 infection, we found that among the COVID-naïve group, 49/111 (44%) had developed breakthrough infections that were either mildly symptomatic (RT-PCR or RAT positive) or asymptomatic (anti-SARS-CoV2 nucleoprotein antibody positive at six months).

Among the COVID-exposed group, 25/94 (27%) vaccinees developed re-infections that were either mildly symptomatic (RT-PCR or RAT positive) or asymptomatic. The breakthrough infections in the COVID-naïve group were significantly higher than the re-infections in the COVID-exposed group (p=0.01; Chi-square test with Yates correction for continuity), suggesting that the natural infection was more efficient in providing subsequent protection than immunization.

Asymptomatic re-infections were determined by a significant increase in anti-spike antibodies at six months after a decline at three months. By three and six months after the second dose, the COVID-naïve group showed a 2-fold and 6-fold decline respectively from the peak antibody levels (one month after the second dose). Similarly, in the COVID-exposed group, the decline was 4-fold and 6-fold at three months and six months respectively from the peak antibody levels (one month after the first dose). Although the peak antibody level time points were different in the COVID-naïve and exposed groups, and were significantly higher in the COVID-exposed group compared with the COVID-naïve group (Figure 1A), the levels declined proportionately in both the groups by six months after the second dose. Thus the marked increase in the antibody levels at three months in the COVID-naïve group with breakthrough infections (13-fold increase) and at six months in the COVID-exposed group with re-infections (1.4-fold increase) are likely to be due to the booster effect from a natural infection, which suggests stimulation of an existing memory B cell pool. The mild or subclinical disease in all the HCWs suggests some protection conferred by the vaccination and / or previous natural infection.

Additionally, we measured the anti-SARS-CoV2 spike antibodies in the serum (Figure 1D) and SWMF (Figure 1E) samples in a small subset of 10 HCWs who took the third booster dose. Among these 10, two were COVID-naïve, two were COVID-exposed prior to the first dose and six belonged to the breakthrough infections group. There was a 7-fold increase in serum antibodies at one month after the third dose (median: 13760 U/ml) compared with six months after the second dose (median: 2054 U/ml). At three and six months after the third dose, there was a marginal 1.3-fold (median: 10900 U/ml) decline and 1.9-fold (median: 5676 U/ml) decline in the serum antibodies. The trend of the rise and decline of the serum antibodies were similar to that of the second dose. Our findings confirm the presence of a memory B cell population that can be activated by specific antigen challenge shown by *in vitro* studies (10). Anti-SARS-CoV2 spike antibodies (assumed to be total Ig including IgG as per manufacturer’s specifications) were detected in all the SWMF samples after the third dose. At one month after the third dose, the median antibody level was 12 U/ml. The antibody levels declined marginally (median: 10 U/ml) at three months and by 2-fold (median: 5 U/ml) at six months. Although the presence of IgG antibodies in the SWMF samples could attribute to protection at the mucosal surfaces, local secretory IgA antibody production was not tested to confirm local mucosal immunity.

The lack of protective immunity from breakthrough infections and re-infections within a short span of less than one year of the antigen/virus exposures raises concerns over a number of host factors that govern a robust protective humoral immunity. These include long lasting antibody titres that are durable and highly avid, isotype changes and protective effect of serum antibody titres at the mucosal surfaces. B cells get activated to elicit a humoral response both by the antigen itself as well as by the antigen-specific T cells. Only a T cell dependent B cell response can provide a long lasting immunity with a robust memory pool. Therefore, in this study we next determined cytokines secreted upon the activation of CD4^+^ helper T cells that are generally involved in the activation and maintenance of B cells. IL-17 is a pro-inflammatory cytokine produced by Th17 cells that also contribute to mucosal immunity. IL-21 activates the Tfh cells that are important in germinal centre formation where B cells get activation signals from antigen-specific T cells. IL-7 and IL-15 are crucial cytokines that maintain the memory T cells required for providing signals to the activated B cells in order to maintain a B cell memory pool.

Of the 192 vaccinees tested for IL-17 levels at one month after the first dose, low levels of IL-17 levels were detected in 106 (55%). COVID-exposed vaccinees elicited significantly higher levels of IL-17 compared to the COVID-naïve group both after the first and second doses of the vaccine (p<0.005, Figure 2A). Similar patterns were found with IL-21; IL-21 levels in 104/190 (55%) vaccinees were low with no significant difference between the COVID-naïve and exposed groups (Figures 2B). The mean IL-7 and IL-15 levels were also only marginally elevated in the vaccinees irrespective of the COVID exposure before or after the vaccination (Figures 2C and 2D). Taken together these findings suggest that both ‘artificial’ immunization as well as a natural infection elicit a poor T cell response during the early inflammatory stage, germinal centre formation for B cell activation and class switching, and maintenance of memory cells. Poor T cell stimulation is probably responsible for the lack of durability and long lasting humoral immune response. Our findings corroborate with the poor germinal centre formation shown at the tissue level in autopsy samples of COVID patients (11). However, studies have shown that T cells and B cells from vaccine primed individuals can be induced *in vitro* by SARS-CoV2 spike peptides (6,7,10), suggesting the presence of memory T cell and B cell pools. Taken together these findings suggest that the primed T cells and B cells have memory, but they are either unable to secrete potent mediators to prevent symptomatic infection or their activity is not reflected in the mucosal surfaces for prevention. At the same time, a quick innate immune response that could clear the virus with minimal recall of the adaptive immune response needs to be evaluated. Effective innate immunity at mucosal surfaces could account for milder COVID-19 disease especially in previously infected / vaccinated individuals.

**Figure 2:**
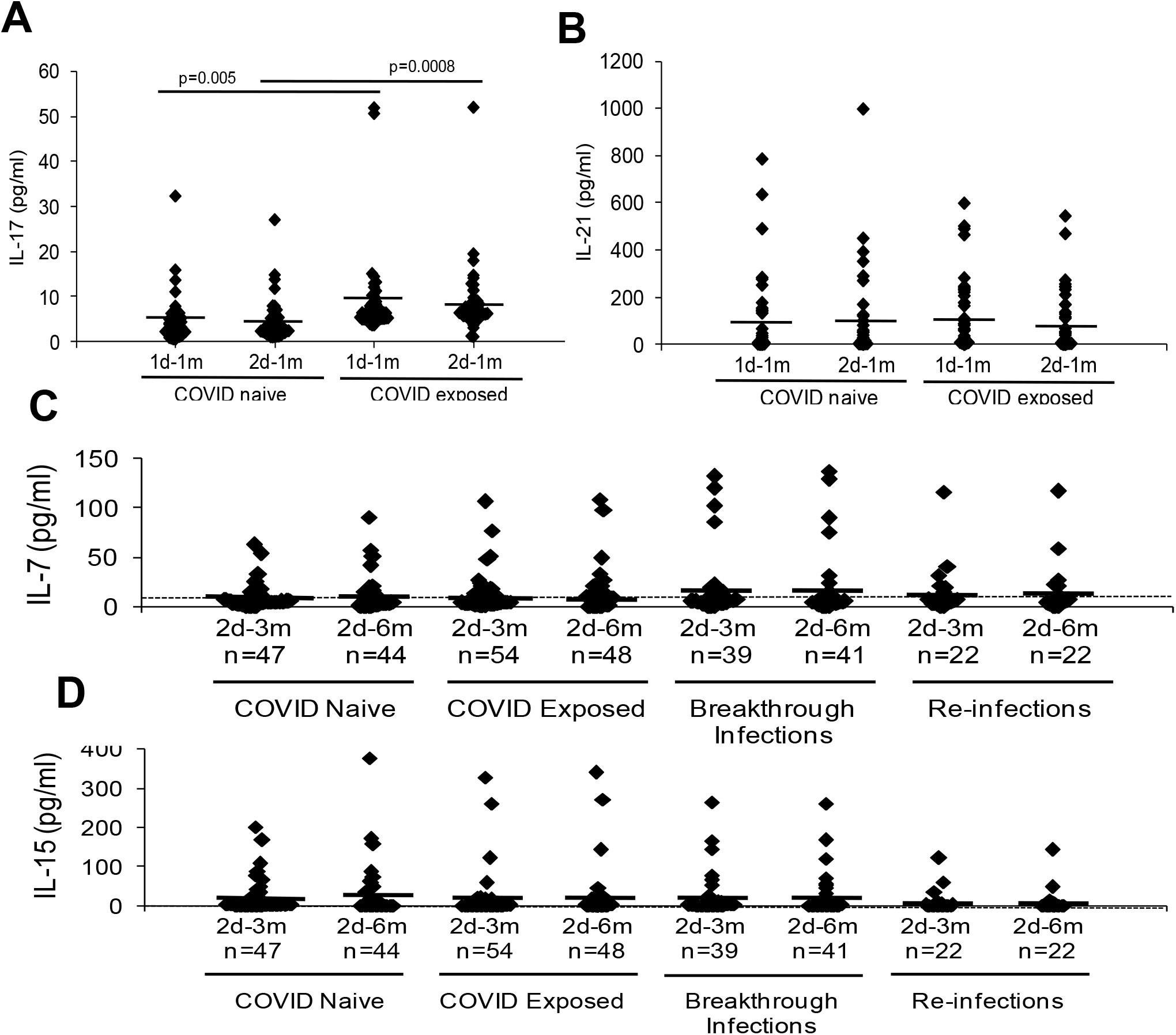
Levels of pro-inflammatory, regulatory and memory cytokines at various time points after vaccination. The X-axis denotes the time points – 1d-1m: one month after the first dose; 2d-1m: one month after the second dose; 2d-3m: three months after the second dose; 2d-6m: six months after the second dose; n denotes the number of samples. Bars show the mean for each group. **A. Levels of IL-17 detected in the COVID-naïve (1d-1m: n=49; 2d-1m: n=52) and COVID-exposed (1d-1m: n=57; 2d-1m: n=56) groups** p values from t-test. **B. Levels of IL-21 detected in the COVID-naïve (1d-1m: n=46; 2d-1m: n-46) and COVID-exposed (1d-1m: n=58; 2d-1m: n=55) groups. C. Levels of IL-7 in COVID-naïve, COVID-exposed, breakthrough infections and re-infections groups at three and six months post-vaccination**. The dotted line depicts serum IL-7 baseline threshold level calculated using the levels of 30 non-responders at 14 days after first vaccine dose. **D. Levels of IL-15 in COVID-naïve, COVID-exposed, breakthrough infections and re-infections groups at three and six months post-vaccination**. The dotted line depicts serum IL-15 baseline threshold levels

Lastly, we analysed the levels of various innate and adaptive immunity mediators in the paired serum and SWMF samples from the 10 HCWs after the third booster dose. Expressions of IL-2, IL-4, IL-12p70, IFN-γ and IL-17 were not detected in either the serum or SWMF samples at both one and six months after the third vaccine dose. Cytokines and chemokines primarily secreted by monocytes and macrophages – MIG, MCP-1, IL-8, TNF-α, IL-6 and IL-1β showed a declining trend in the serum samples, but an increasing trend in the SWMF samples between the first month and the sixth month time points after the third dose (Figure 3). Although IL-10 and IL-21 also showed a similar trend, the detection rate and levels are quite marginal (data not shown). Taken together, this preliminary study on a small subset of 10 HCWs suggest a local innate immune response involvement, which lasts longer in the oral mucosa than the periphery. Considering that SWMF is swallowed and renewed, this suggests the continuing presence of local active immune processes.

**Figure 3:**
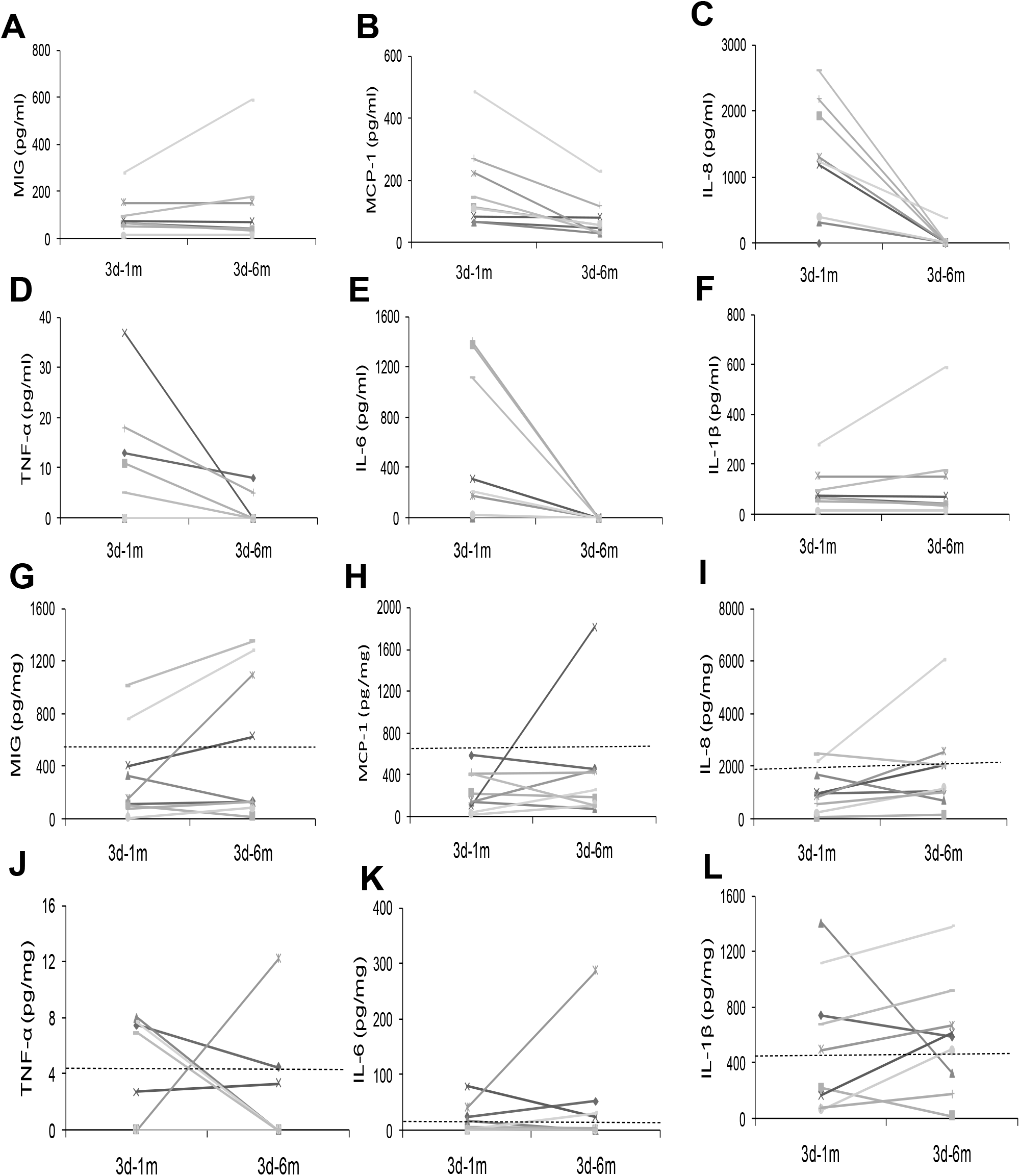
Comparison of cytokines and chemokines in serum (A to F) and SWMF (G to L) in 10 HCWs one and six months after the third vaccine dose. Concentration of the cytokines/chemokines in serum (pg/ml) and SWMF samples (pg/mg of total protein). Dotted lines (G to L) in SWMF samples denote the baseline levels of the corresponding cytokines/chemokines generated using SWMF samples from 11 unvaccinated COVID-naïve controls. **Monokine in response to interferon gamma (MIG) in serum (A) and SWMF samples (G). Monocyte chemoattractant protein-1 (MCP-1) in serum (B) and SWMF samples (H). IL-8 in serum (C) and SWMF samples (I). Tumour necrosis factor–alpha (TNF-α) in serum (D) and SWMF samples (J). IL-6 in serum (E) and SWMF samples (K). IL-1β in serum (F) and SWMF samples (L)**.

In conclusion, our study shows that the Covishield vaccine, an adenoviral vector-based vaccine carrying the DNA coding for the spike protein elicits a significant antibody response after the first and second doses. The antibody response follows a prime-boost pattern in COVID-naïve individuals. In COVID-exposed individuals, the first dose acts as a boost to the antibody response, while the second dose appears to induce antibody anergy. The presence of antibodies provides protection from disease severity and minimizes hospitalization; it does not appear to prevent acquisition of a mild or asymptomatic re-infection. This may be due to viral factors like mutations of the structural proteins or host factors like changes in the titre, avidity or isotype of the antibodies, or that serum antibodies are not present in sufficient amounts at the mucosal sites of infection. The T cell responses required to activate and maintain a robust antigen-specific B cell pool have been minimally elicited both by the vaccine as well as the natural infection as indicated by the low levels of IL-17, IL-21, IL-7 and IL-15 in all groups of vaccinees – COVID-naïve, COVID-exposed, breakthrough infections and re-infections. The minimal T cell response suggests that the humoral immunity elicited by the vaccine/natural infection will potentially be short-lived with limited memory, thereby requiring frequent boosters in order to maintain circulating antibodies that confer protection against disease severity. Additionally, a robust innate immune response leading to early virus clearance is elicited and is long-lasting in the oral mucosa. Further in-depth explorations of the mucosal innate immune response at various stages of the disease are warranted.

## Data Availability

All data produced in the present work are contained in the manuscript.

